# FMT Restores Colonic Protein Biosynthesis and Cell Proliferation in Patients with Recurrent *Clostridioides difficile* Disease

**DOI:** 10.1101/2024.11.28.24318101

**Authors:** G. Brett Moreau, Mary Young, Brian Behm, Mehmet Tanyüksel, Girija Ramakrishnan, William A. Petri

## Abstract

Recurrent *C. difficile* infection (CDI) is a major health threat with significant mortality and financial costs. Fecal Microbiota Transplantation (FMT) is an effective therapy, however the mechanisms by which it acts, particularly on the host, are poorly understood. Here we enrolled a prospective cohort of human patients with recurrent CDI (n=16) undergoing FMT therapy. Colonic biopsies were collected and bulk RNA sequencing was performed to compare changes in host gene expression pre- and two months post-FMT. Transcriptional profiles were significantly altered after FMT therapy, with many differentially expressed genes (∼15% of annotated genes detected). Enrichment analysis determined that these changes were reflective of increased protein production post-FMT, with enrichment of pathways such as Ribosome Biogenesis, Protein Processing, and signaling pathways (Myc, mTORc1, E2F) associated with cell proliferation and protein biosynthesis. Histology of H&E-stained biopsies identified a significant increase in colonic crypt length post-FMT, suggesting that this treatment promotes cell proliferation. Crypt length was significantly correlated with enriched Myc and mTOR signaling pathways as well as genes associated with polyamine biosynthesis, providing a potential mechanism through which this may occur. Finally, signaling pathways upstream of Myc and mTOR, notably IL-33 Signaling and EGFR ligands, were significantly upregulated, suggesting that FMT may utilize these signals to promote cell proliferation and restoration of the intestine.

## Introduction

*Clostridioides difficile* infection (CDI) is a critical public health threat that is associated with significant morbidity, mortality, and financial costs. Although the CDI-associated mortality rate in the United States is 2.7% for primary infection, there is a 20% recurrence rate (1), and mortality is nearly 10 times higher (25.4%) for recurrent disease (2). While standard of care for CDI remains antibiotic treatment (3, 4), antibiotic exposure is a significant risk factor for recurrent CDI (5) due to its disruption of the intestinal microbiota. Because of this, fecal microbiota transplantation (FMT) has emerged as a major therapeutic option for recurrent CDI. Two FMT therapeutics have recently been FDA-approved for the treatment of recurrent *C. difficile*: SER-109 (Vowst), an oral therapeutic utilizing purified *Firmicutes* spores (6) and RBX2660 (Rebyota), which uses a consortium of live microbes and is delivered via enema (7). As both products are of limited efficacy, better understanding of the specific mechanisms underlying FMT offer the promise for improved treatment.

Our understanding of the mechanisms through which FMT is protective, particularly its effect on the host, remain incomplete. Because CDI is associated with antibiotics and microbiome disruption (8, 9), much of the work looking at FMT’s role in recurrent CDI has focused its impact on restoration of the intestinal microbiota (10) and resulting microbial metabolites, such as bile acids (11–13). These results suggest that FMT may protect against *C. difficile* through niche restriction and competition with the transplanted microbes (14). However, signaling from the microbiota can also affect intestinal immune cell development and signaling (15), and it is likely that FMT also initiates changes in the intestinal epithelium or immune cells that impacts successful protection from recurrent disease. A role for the immune response in the pathogenesis of primary CDI has previously been identified, as elevated immune responses and increased inflammation in response to toxin-mediated tissue damage are associated with worse clinical outcomes independently of bacterial burden (16–18). In addition, the type of immune response can have a significant impact on disease progression, as Type 3 immune responses have been associated with exacerbated disease severity (19), while Type 2 immunity has been associated with tissue repair and protection from severe disease (20–22). Based on these data, we hypothesized that FMT directly impacts host responses in the gut to promote protection against recurrent CDI.

We have recently identified gene expression changes associated with the initial stages of FMT in a mouse model of antibiotic treatment (23). This study found significant changes in host immune responses both transcriptionally and in immune cell populations within one week of FMT administration. FMT was also associated with upregulation of genes associated with intestinal homeostasis and neuropeptide signaling, which suggest that FMT can promote restoration of intestinal homeostasis in the absence of prior *C. difficile* exposure. While the effects of FMT on the host have become better understood, many of these studies have looked in the context of other inflammatory disorders, such as intestinal colitis (24–26). In addition, data in patient cohorts is still limited, as most studies have utilized animal models. Many studies with FMT patients have been limited to the assessment of systemic biomarkers, finding that FMT can alter inflammatory biomarkers (27, 28) as well as miRNA profiles, which can impact immune signaling (29). Both immune and colonic transcriptional changes have previously been characterized by our group in a cohort of patients receiving FMT for recurrent CDI (30). This study observed increased expression of the Type 2 cytokine IL-25 along with increased expression of genes promoting intestinal epithelial cell differentiation and extracellular matrix restoration post-FMT. However, this study was limited by a small cohort size (n=6) and the confounder of IBS-like symptoms in two patients in the study.

The current study aimed to replicate these findings in a larger patient cohort with a more expansive collection of biological samples, allowing for more thorough characterization of the biological changes associated with FMT. We utilized bulk RNA sequencing of colonic biopsies pre- and two months post-FMT to investigate FMT-driven changes in host transcriptional profiles. Our results suggest that FMT promotes expression of protein biosynthesis and extracellular matrix remodeling pathways, potentially through upstream IL-33 and EGFR signaling. These changes may promote increased cell proliferation within the colon and restoration of intestinal homeostasis.

## Methods

### Study Enrollment

Participants in this study were drawn from an ongoing clinical study at the University of Virginia. This study has been approved by the Institutional Review Board and is registered under ClinicalTrials.gov ID NCT02797288. Study subjects (aged 18-85) were recruited from recurrent CDI patients scheduled for FMT therapy through colonoscopy in the UVA outpatient clinic. Donor FMT material was obtained from a screened stool bank (OpenBiome, Cambridge, MA). Follow-up collection of biopsies was also performed approximately two months (mean = 63.2 days) from the date of FMT administration.

### Biopsy Collection and Preservation

Biopsy samples were collected from the distal colon during the FMT colonoscopy and by sigmoid colonoscopy at scheduled follow-up. Biopsies were either immediately flash frozen or stored in Allprotect tissue reagent (Qiagen) and then flash frozen. Frozen biopsies were then stored at −80°C until analysis. Two biopsies were used at both pre- and post-FMT timepoints for RNA isolation and subsequent sequencing. Due to a change in collection protocol, two Allprotect biopsies were used for six patients, while one AllProtect biopsy and one flash frozen biopsy were used for the other ten patients. Additional biopsies were formalin-fixed and paraffin-embedded (FFPE) for Hematoxylin and Eoxin (H&E) staining and tissue visualization.

### Bulk RNA Sequencing and Bioinformatics

Biopsies stored for RNA sequencing as described above were first transferred to clean 2ml screw-cap tubes containing a sterile 5mm stainless steel bead, then homogenized using a TissueLyser II (Qiagen) for 3 minutes at 25Hz. Homogenized tissue was vortexed and then RNA isolated using the RNeasy kit (Qiagen) according to the manufacturer’s protocol. RNA yield and quality was evaluated using a TapeStation (Agilent), then stored at −80°C until use. Purified total RNA was submitted to Novogene for bulk RNA sequencing. Libraries were generated after Poly(A) enrichment for mRNA transcripts, followed by paired-end 150 base pair sequencing using a NovaSeq X Plus series sequencer (Illumina).

Prior to analysis, unprocessed sequencing reads were evaluated for quality using FastQC (31) and MultiQC (32), trimmed to remove adapter sequences using BBTools, and pseudomapped to the human genome using Kallisto (33). The resulting count tables were imported into R (34) using TxImport (35) and the DESeq2 package (36) was used to exclude genes with low counts, normalize data, estimate dispersions, and fit counts using a negative binomial model. Differential gene expression was determined based on this multivariate model.

Gene Set Enrichment Analysis (GSEA) was performed using the fgsea package (37) in R. Briefly, all genes included in the data set were ranked from most upregulated to most downregulated post-FMT according to their Wald statistic from the multivariate model. This ranked list was used for GSEA using the Hallmark (38), Gene Ontology (39), or Kyoto Encyclopedia of Genes and Genomes (40) data sets. The tidyverse package (41) was used for data organization and visualization.

### Histology and Crypt Length Measurement

Slides containing FFPE biopsy sections were stained with H&E and imaged for crypt length quantification. At least three crypts from two different regions were selected from each slide. Crypt lengths were calculated using Sedeen slide viewer software (Pathcore). Researchers were blinded to the FMT status of each slide at the time of histopathologic analysis. After collection of all scores, means and standard deviations were calculated on a per-slide and per-patient basis at both pre-FMT and post-FMT timepoints. Statistics were performed on per-patient average pre- and post-FMT crypt lengths using a linear mixed effects model that incorporated patient ID to control for within-patient variability.

## Results and Discussion

### Patient Characteristics

A total of 16 patients with recurrent CDI undergoing FMT therapy provided colonic biopsy samples at both FMT and follow-up appointments. The clinical characteristics of this cohort are summarized in Table 1. Patients in the cohort were majority female and entirely white. While 94% of patients had at least 3 recurrences prior to FMT (mean = 3.8), FMT therapy was successful in all patients as defined by no recurrences within the follow-up window. All patients were treated with vancomycin prior to FMT treatment (standard of care at UVA hospital), and no patients used antibiotics during the follow-up period post-FMT. IBD was not a major confounder of this study, as only one patient had an IBD diagnosis in this cohort.

**Table 1:** Clinical Characteristics of the FMT Cohort.

| Clinical Characteristic |  | FMT Cohort (n=16)* |
| --- | --- | --- |
| Mean age at FMT |  | 65 (12.1) |
| Number male gender |  | 6 (37.5%) |
| Number white ethnicity |  | 16 (100%) |
| Mean BMI at FMT |  | 29.9 (6.1) |
| Mean number of CDI recurrences prior to FMT |  | 3.8 (1.1) |
| Number of patients with: | 2 recurrences | 1 (6.25%) |
|  | 3 recurrences | 6 (37.5%) |
|  | 4 recurrences | 6 (37.5%) |
|  | 5 recurrences | 1 (6.25%) |
|  | 6 recurrences | 2 (12.5%) |
| Number patients treated with vancomycin prior to FMT |  | 16 (100%) |
| Number of patients with IBD |  | 1 (6.25%) |
| Number of patients with IBD Subtypes | Crohn's Disease | 1 (6.25%) |
|  | Ulcerative Colitis | 0 (0.0%) |
| FMT Success Rate (No recurrences within 60 day follow-up) |  | 16 (100%) |
| Number of patients with current or prior antibiotic use at follow-up. |  | 0 (0.0%) |

\*Represents Mean (Standard Deviation) or Total Number (Percentage)

### FMT Promotes Broad Changes in Intestine Transcriptional Profiles

To evaluate the effect of FMT on host gene expression, bulk RNA sequencing (RNAseq) was performed on biopsies collected immediately pre-FMT and at two month follow-up. Reads from bulk RNAseq were processed to preserve only high quality reads (>Q30, indicating 99.9% base calling accuracy) and remove adapter content. After this, the clean reads were pseudomapped to the human genome using Kallisto, with 80% of these reads successfully mapped and used for downstream analysis. Differentially Expressed Genes (DEGs) were calculated using a negative binomial multivariate model that incorporated patient ID number to account for within-patient variability. A total of 1,877 genes were significantly upregulated post-FMT (adjusted p value < 0.05), while 1,788 were downregulated (Fig. 1A). The 3,665 total DEGs represented approximately 15% of annotated genes included in the analysis. Of these differentially expressed genes, 154 (8.2%) and 182 (10.2%) genes had at least a two-fold increase or decrease in gene expression, respectively. Together, these results indicate that FMT promotes large changes in gene expression that persist out to two month follow-up.

**Figure 1:**
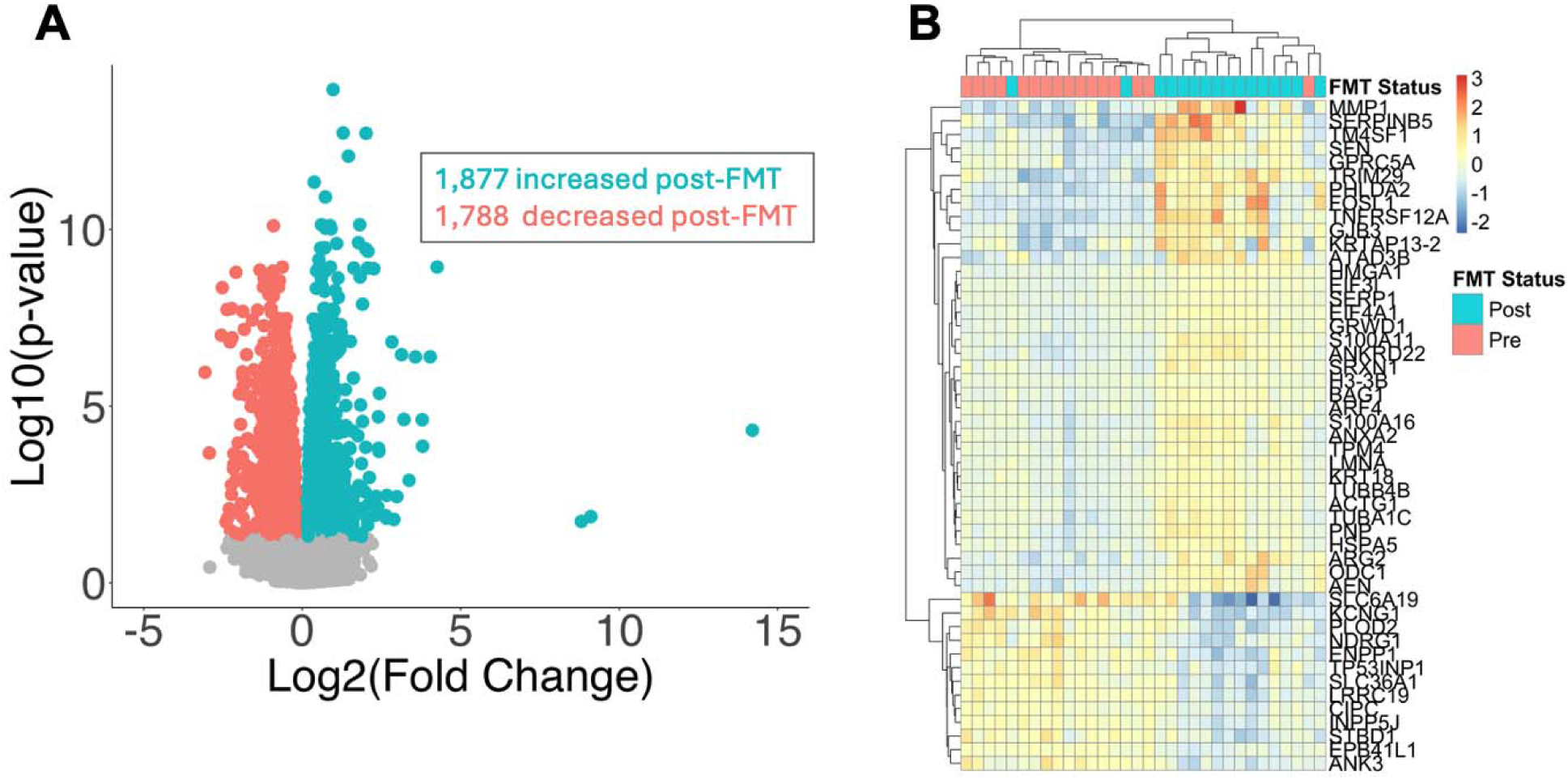
FMT drives changes in host transcriptional profiles. Colonic biopsy samples were collected immediately pre-FMT or at two month follow-up post-FMT and used to evaluate host transcriptional profiles using bulk RNA sequencing. A) Volcano plot showing differentially expressed genes (DEGs) that are increased (blue) or decreased (red) in patients post-FMT compared to baseline. B) Heatmap of 50 most differentially expressed genes between pre- and post-FMT biopsy samples.

The 50 most differentially expressed genes between timepoints are presented in Figure 1B. Hierarchical clustering was performed with these data and samples clustered primarily according to FMT status. The most prominent gene significantly increased post-FMT was *FOSL1*, a FOS family protein that heterodimerizes to form the transcription factor AP-1 (42). *FOSL1* is upregulated in cancer cells in a kRAS-dependent manner (43) and is associated with increased stemness in cancer cells, potentially through mutual regulation with NF-kB (44). Other significantly increased genes included genes associated with extracellular matrix remodeling, such as *MMP1* and *SERPINB5*. These genes have been implicated as markers of epithelial-mesenchymal transition and poor prognosis in colorectal cancer samples (45). Downregulated genes include genes such as *SLC6A19*, *KCNG1*, *ENPP1*, *NDRG1*, and *PLOD2*. SLC6A19 is a neutral amino acid transporter that is strongly repressed in stem cells via SOX9 (46), and loss of this gene is associated with protein restriction and decreased mTORc1 activity (47). ENPP1 is an extracellular cGAMP hydrolase, and its activity is associated with degradation of immunomodulatory signaling and less efficient inhibition of cancer growth and metastasis (48). Overall, these results point to a more proliferative environment post-FMT.

These findings were broadly consistent with findings from our previous cohort: 51.6% of differentially expressed genes in Jan *et al* were also significantly altered in this data set. This includes several of the most differentially expressed genes from Figure 1B, such as *MMP1*, *SERPINB5*, and *SLC6A19*.

### Pathway Analysis with RNAseq Data

We aimed to take a more systematic approach to identifying biological pathways that were altered by FMT. To do this, RNA sequencing data were analyzed using Gene Set Enrichment Analysis (GSEA) to identify potential functions that were enriched pre- or post-FMT. Three different databases were used for this analysis: Hallmark, Gene Ontology: Biological Processes (GO:BP) and Kyoto Encyclopedia of Genes and Genomes (KEGG). We took this approach to increase our coverage of biological pathways, as each database contains slightly different gene sets and together they provide a more comprehensive view of pathways whose expression may be modified by FMT. The top five enriched gene sets for each database in both upregulated and downregulated post-FMT conditions are presented in Figure 2, while all gene sets are presented in Supplementary Table 1.

**Figure 2:**
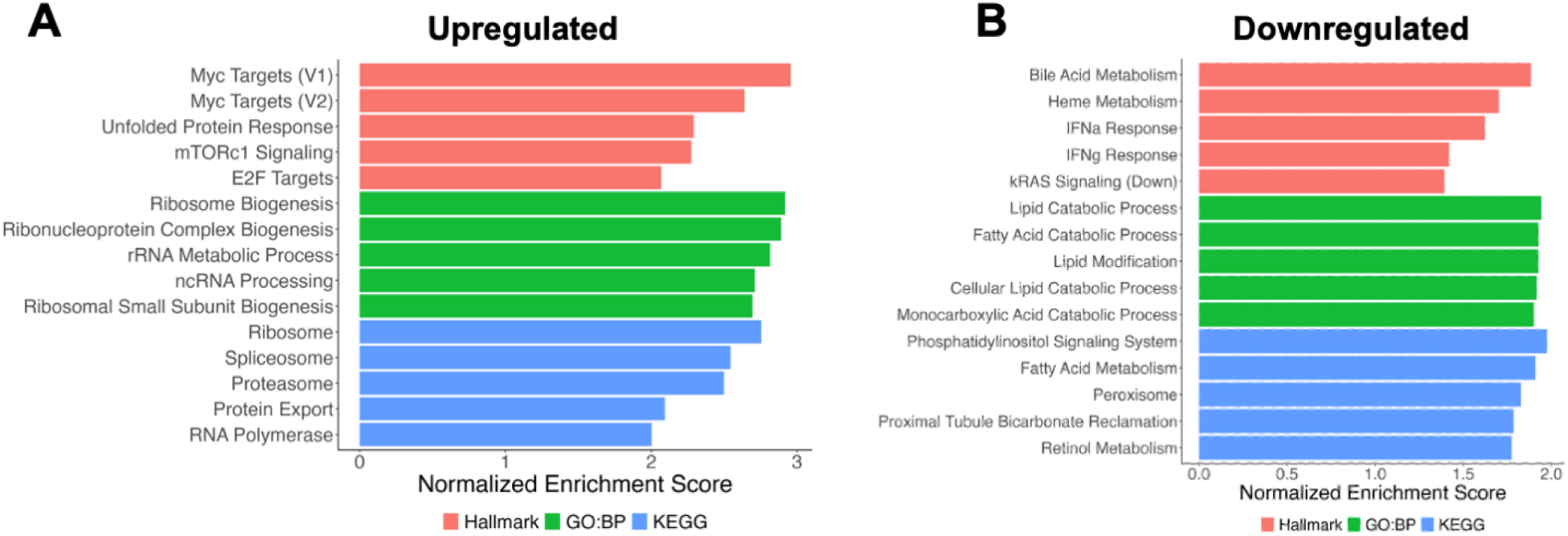
Gene sets associated with ribosome/protein synthesis and lipid metabolism are altered by FMT. Ranked barplot of the most significantly increased (A) or decreased (B) gene sets post-FMT as determined by Gene Set Enrichment Analysis (GSEA). Top five gene sets are shown for the Hallmark (red), Gene Ontology: Biological Processes (green) and KEGG (blue) databases. Genes are ranked according to Normalized Enrichment Score.

The most prominently upregulated process in post-FMT biopsy samples was ribosomal biogenesis and protein processing (Fig. 2A). Ribosome and protein synthesis gene sets were significantly enriched post-FMT in both GO:BP and KEGG databases, making up most of the top 5 enriched gene sets. Analysis of leading edge genes (those driving enrichment of each gene set in GSEA) found that GO:BP gene sets were primarily driven by the same core set of genes. These genes were primarily regulators of ribosome biogenesis and maturation (*RRP15*, *RRS1*, *BYSL*, *UTP11*, *MAK16*) or nuclear proteins (*NUP88*, *NOP16*), many of which have been implicated in cell proliferation in models of cancer (49–51). Ribosome biosynthesis, a process that starts in the nucleus and continues in the cytoplasm, is a critical process required for the level of protein production necessary for cell proliferation (52, 53). GSEA using KEGG gene sets also identified Ribosomes as the most strongly enriched gene set along with other gene sets associated with gene expression (Spliceosome, RNA Polymerase) and subsequent protein processing (Protein Export, Proteasome). This highlights the high level of metabolic activity post-FMT through the synthesis of new proteins.

Hallmark gene sets exhibited slightly different gene sets. The most enriched gene sets from this database were downstream targets of several signal transduction molecules, including Myc, mTORc1, and E2F. Myc and mTORc1 are major regulators of ribosomal biogenesis and subsequent protein synthesis (54, 55), and many of the genes observed in these gene sets are indicative of these processes. The Unfolded Protein Response was also upregulated post-FMT, including genes such as *HYOU1*, *HSPA4*, and *BAG3*, which are associated with response to misfolded protein stress that may result from increased protein synthesis. Similarly, Proteosome genes were also enriched in the KEGG database and are likely a response to increased protein synthesis and turnover. Proteasome machinery is critical for proper cell proliferation, and proteasome inhibitors are a clinically approved therapy for several cancers (56).

Several gene sets were downregulated post-FMT (Fig. 2B). The most prominent downregulated signature was fatty acid catabolism, which was prominent in the Top 5 most enriched gene sets for both GO:BP and KEGG databases. This was driven by enzymes in the fatty acid beta-oxidation pathway, including *ACOX1, ACAA2, ADH1C,* and *ACSL5*. Bile acid metabolism was also significantly decreased post-FMT. These changes in lipid metabolism, namely downregulation of fatty acid and bile acid metabolism pathways, were also observed post-FMT in our previous study investigating a mouse model of FMT treatment (57). The microbiota play a significant role in both lipid metabolism (58) and insulin sensitivity (59), so these transcriptional changes may be due to differences in host metabolism and energy production driven by differences in microbial metabolism of the diet. Indeed, host energy metabolism pathways Glycolysis and Oxidative Phosphorylation were both significantly enriched post-FMT (Suplementary Table 1), suggesting that a shift from Glycolysis to fatty acid beta-oxidation may occur in the absence of the microbiome. Bile acids also play a major role in lipid absorption through fat emulsification and solubilization (60). The intestinal microbiota are capable of modifying these bile acids and antibiotic treatment disrupts this process, altering intestinal bile acid composition (61).

IFN and IFN signaling were also decreased post-FMT, and these pathways were primarily driven by a core set of interferon-stimulated (*IFIT1*, *IFIT2*, *GZMA*) and apoptosis-associated (*CASP7*, *CASP3*) genes. Gene sets associated with Bile Acid Metabolism, Fatty Acid Metabolism, and IFN and IFN Responses were all enriched in antibiotic-treated mice compared to naive controls (62), indicating that these transcriptional changes are likely due to antibiotic-mediated disruption of the intestinal microbiota. Finally, genes that were downregulated by kRAS signaling were also downregulated post-FMT, which is consistent with the upregulation of *FOSL1*, a kRAS-upregulated gene (43), observed in Figure 1B. kRAS is a small GTPase that plays a key role in maintenance of intestinal homeostasis. It is activated by many extracellular stimuli, including EGFR signaling, and is able to transduce those signals through activation of downstream signaling pathways such as Raf/Erk and PI3K/Akt/mTOR to promote cell proliferation or intestinal renewal (63). This served as additional evidence that FMT promotes signaling pathways that promote cell proliferation.

### FMT is associated with Colonic Crypt Elongation that Correlates with Myc and mTORc1 Target Gene Expression

Because we observed enrichment of gene sets associated with increased metabolic activity (Ribosome Biogenesis, Protein Export, Myc and mTORc1 Signaling), we hypothesized that the epithelium would experience significant proliferation post-FMT. To test this, we measured crypt length in H&E stained colonic biopsies, as crypt elongation has been associated with elevation of cell proliferation markers such as Ki67 or LGR5 (64, 65). Crypt length was measured from multiple slides per patient at each timepoint in a blinded fashion. Statistical differences were determined by comparing the average crypt length pre- and post-FMT in a linear model controlling for within-patient variability. Average crypt length was significantly longer in patients post-FMT (Fig. 3A-B), suggesting that FMT stimulated epithelial cell proliferation. Of note, CDI was previously associated in a mouse study with elongated crypt length, with FMT paradoxically restoring colonic crypts to a shorter length more similar to uninfected mice (66). This finding was in the context of acute infection with *C. difficile* and the resulting inflammatory immune response, which was already promoting increased crypt length. This is consistent with other infections where crypt elongation has been observed, such as *Salmonella* Typhimurium (67) and HIV (68). Broad inflammatory signaling was largely absent from our RNA sequencing data, consistent with our earlier finding that FMT promotes Type 2 immunity in humans (30). We hypothesize that this anti-inflammatory colonic environment likely contributes to the differential effects of FMT on crypt length.

**Figure 3:**
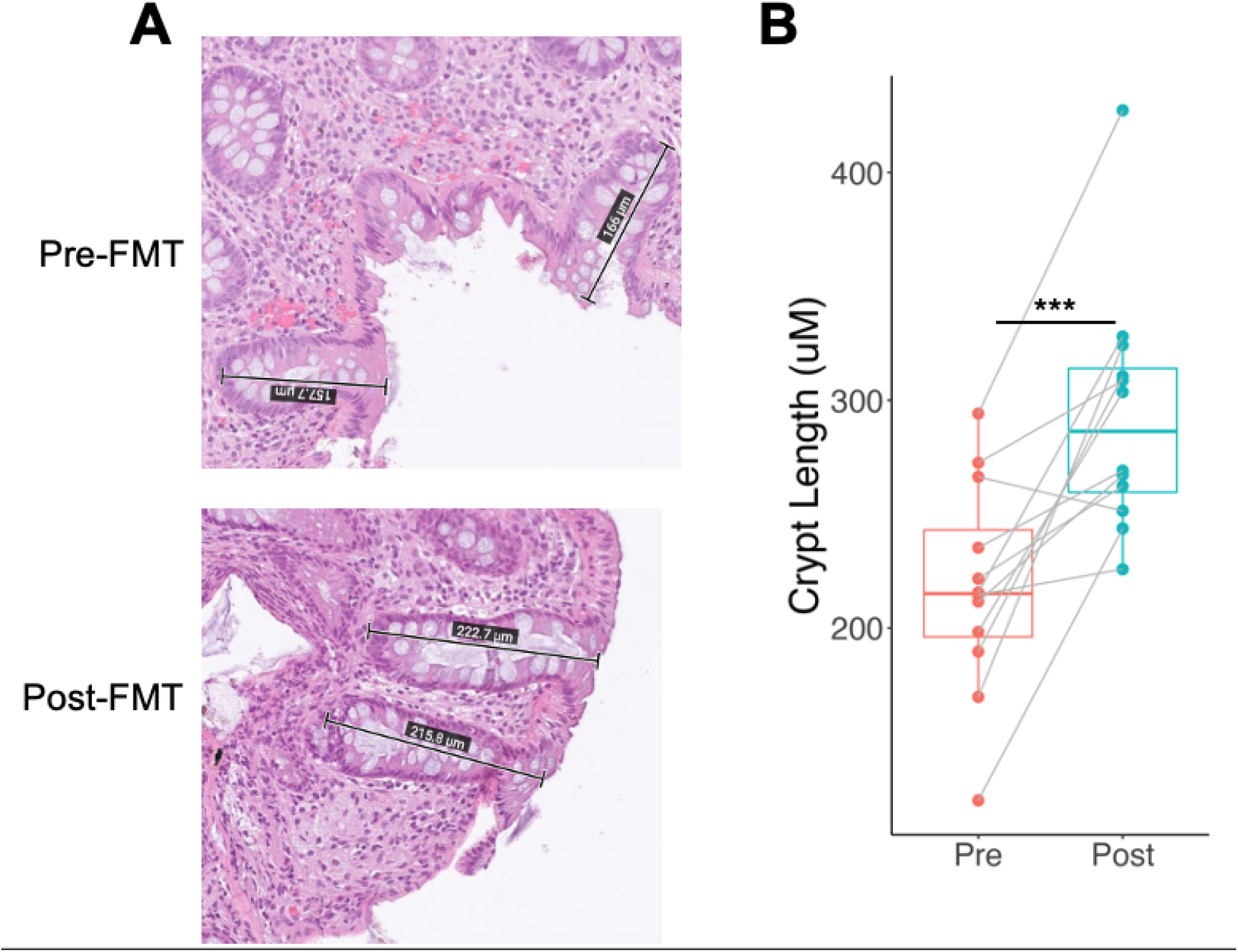
FMT promotes increased colonic crypt length. A) Representative H&E images of crypts from colonic biopsies from the same patient pre- and post-FMT. B) Quantification of average crypt length across all patients. Each dot represents a single patient and gray lines connect paired samples. Statistics were calculated using a linear mixed effects model controlling for within-patient variability. ***, p < 0.001

One of the strongest signatures from the RNAseq data was Myc target genes. Because these signaling pathways are known to promote ribosome biogenesis, protein production, and cell proliferation, we hypothesized that crypt length would correlate with the expression of target genes in these pathways. To test this, expression of the top 10 leading edge genes for both Myc and mTORc1 Target Genes were tested for correlation with crypt length from both pre- and post-FMT samples. Overall, 6/10 Myc Target genes and 4/10 mTORc1 Target leading edge genes were significantly correlated with crypt length, and all trended towards a positive correlation (Fig. 4A), indicating that these pathways are associated with increased crypt cell proliferation. Previous work has shown the Myc is required for proliferation of crypt progenitor cells, as crypts in which Myc has been knocked out are quickly lost and replaced by Myc-sufficient crypts (69). These Myc-deficient crypts were associated with fewer cell numbers per crypt, smaller cell sizes, and reduced biosynthetic activity compared to Myc-sufficient crypts. The most differentially expressed Myc target gene in our data set was *ODC1*, the rate limiting enzyme for polyamine biosynthesis. *SRM*, another polyamine biosynthesis gene, was also a Myc Target gene, further implicating this pathway. Polyamine biosynthesis genes, including *ODC1*, have been implicated in ribosome homeostasis, as depletion of these genes was associated with impaired biogenesis (70). Polyamines are also important sources of energy for enterocytes, making them critical for epithelial renewal and proper barrier function in the gut (71), which supports their potential role in cell proliferation. Both *ODC1* and *SRM* expression were significantly correlated with colonic crypt length (Fig. 4B-C), suggesting that polyamine biosynthesis and utilization may be one potential mechanism by which crypt elongation occurs. Overall, these results provide additional evidence that FMT treatment promotes cell proliferation in the intestinal epithelium.

**Figure 4:**
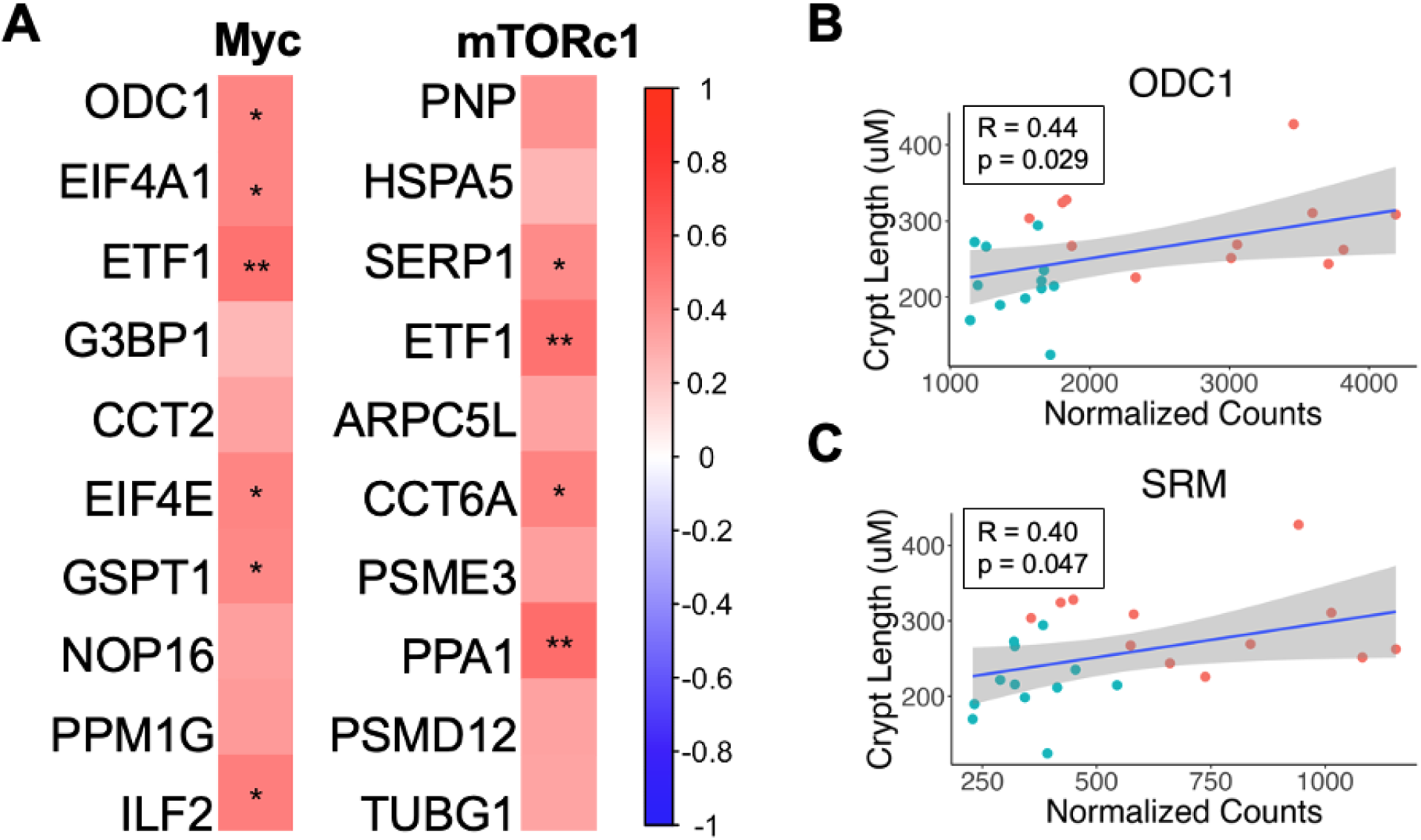
Crypt length is correlated with Myc and mTORc1 target genes. A) Heatmap showing the Pearson correlation R value for the top 10 leading edge genes driving enrichment of Myc and mTORc1 Target gene sets in GSEA. Significantly correlated genes are indicated with asterisks. B-C) Correlation plots showing correlation between average crypt length and normalized gene counts for *ODC1* (B) or *SRM* (C), rate limiting genes for polyamine biosynthesis. *, p < 0.05; **, p < 0.01

### IL-33 and EGFR Signaling Ligands Are Upregulated Post-FMT

Based on the enrichment in Myc and mTORc1 target genes post-FMT and their association with increased protein production and changes in crypt length, we were interested in determining potential upstream mechanisms of action. mTORc1 and E2F signaling cascades can both be stimulated by EGFR signaling through PI3K/Akt and Ras/Raf/Erk signaling cascades, respectively (72). In addition, Ras/Raf/Erk and PI3K/Akt signaling can significantly increase the half-life of Myc protein and thus enhance its effects on transcription (73). Based on these data, we hypothesized that EGFR signaling may be partially responsible the significant increase in Myc, mTORc1, and E2F target genes observed in our GSEA. We observed that several ligands of EGFR were significantly upregulated post-FMT, including amphiregulin (*AREG*), epiregulin (*EREG*) and heparin-binding EGF-like growth factor (*HB-EGF*) (Fig. 5A).

**Figure 5:**
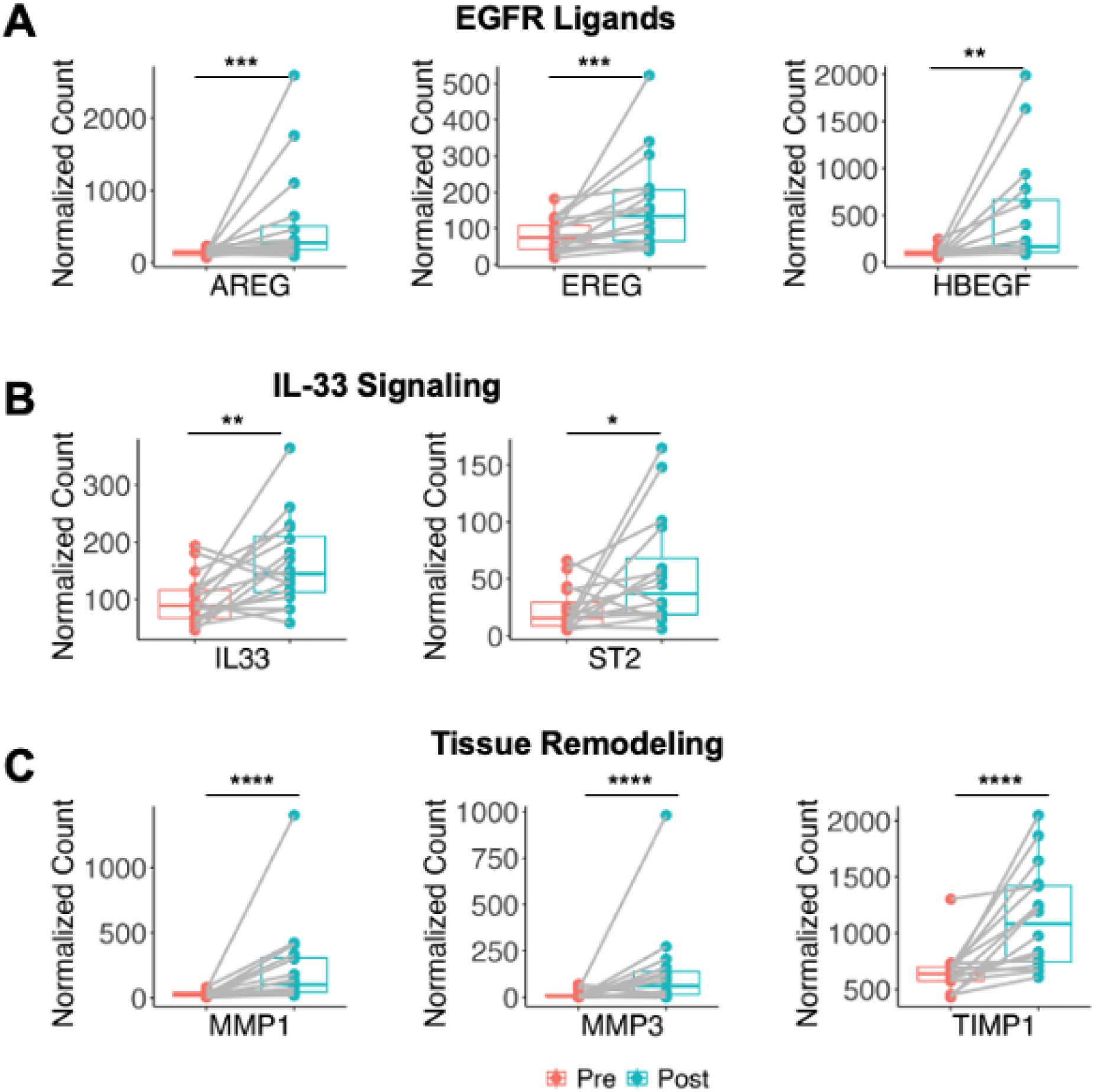
FMT promotes expression of IL-33 signaling, EGFR ligand, and tissue remodeling genes. Normalized gene counts of select A) EGFR ligand, B) IL-33 signaling, and C) tissue remodeling genes either pre-FMT (red) or post-FMT (blue). Statistics are derived from the DESeq2 multivariate model, which adjusts for multiple comparisons. *, p < 0.05; **, p < 0.01, ***, p < 0.001; ****, p < 0.0001

Previous studies in our group have identified Type 2 immune responses in both animal models of FMT and human patients (23, 30). Innate Lymphoid Cell 2s (ILC2s), which are critical for the intestinal Type 2 immune response, are responsive to signals from the microbiome (74). Our group has recently observed that antibiotic treatment significantly decreased ILC2 populations, while IL-33 treatment restored these populations (75). ILC2s robustly produced AREG in response to IL-33 treatment, and this was protective against acute CDI. Based on these data, we hypothesized that Type 2 immunity, particularly IL-33 signaling, may be upregulated post-FMT. We observed that IL-33 signaling was differentially expressed, with transcripts of both the cytokine (*Il33*) and receptor (*IL1RL1*/ST2) significantly increased post-FMT (Fig. 5B). ILC2-derived AREG can signal through EGFR to promote intestinal tissue repair, and disruption of this signaling is associated with inflammatory bowel disease in both animal and human models (76). In addition, HB-EGF signaling through EGFR can feed back on this process to increase IL-33 expression, and this upregulation is required for wound repair in HB-EGF treated keratinocytes (77). IL-33/ST2 Signaling and resulting production of AREG and other EGFR ligands may therefore promote cell proliferation and intestinal repair observed post-FMT.

IL-33 signaling has been shown to promote expression of several matrix metalloproteinases (MMPs) (78), which may be one mechanism by which FMT promotes tissue remodeling and restoration of homeostasis. We identified extracellular matrix remodeling genes (*MMP1*, *SERPINB5*) as some of the most highly upregulated genes post-FMT (Fig. 1B). Consistent with this, we also found that the gene set for epithelial-mesenchymal transition (EMT), which is associated with tissue repair and regeneration, was significantly enriched post-FMT by GSEA (Supplementary Table 1). EMT is characterized by the transition of polarized epithelial cells to mesenchymal cells with increased cell mobility and increased production of extracellular matrix proteins. As part of this process, these cells also upregulate MMPs, which can remodel the extracellular matrix, along with MMP inhibitors such as tissue inhibitors of metalloproteases (TIMPs) or serine protease inhibitors (SERPINs) (79). Further targeted analysis of differentially expressed genes identified several MMPs that were significantly increased post-FMT (Fig. 5C) as well as inhibitors of this process (*TIMP1*, *SERPINB5*). MMP regulation can be induced by several signaling cascades, including NF-kB and MAPK pathways as well as AP-1 binding to MMP promoter regions (80), all pathways that have been implicated in our data. Furthermore, knockdown of MMP1 using shRNA resulted in decreased EMT and decreased Akt and Myc expression in a colorectal cancer cell line (81), suggesting that expression of these genes may sustain activation of these pathways. Overall, these results are consistent with an environment associated with intestinal regeneration and repair.

## Conclusions

A potential model of how the pathways implicated by our transcriptional data may contribute to changes in crypt length and intestinal repair is presented in Figure 6. We observed that post-FMT colonic biopsies exhibit longer crypt length than those pre-FMT, suggesting that FMT promotes cell proliferation. Utilizing bulk RNA sequencing of these biopsy samples, we observed distinct expression profiles between conditions with many differentially expressed genes, particularly in pathways associated with protein synthesis and processing as well as extracellular matrix reorganization. Target genes in the Myc and mTORc1 pathways were significantly correlated with crypt length in these patients, including genes associated with polyamine biosynthesis, an important pathway for energy production in enterocytes. We also observed significant changes in IL-33 signaling and EGFR ligand genes, suggesting that IL-33 signaling and resulting production of EGFR ligands may underlie these changes.

**Figure 6:**
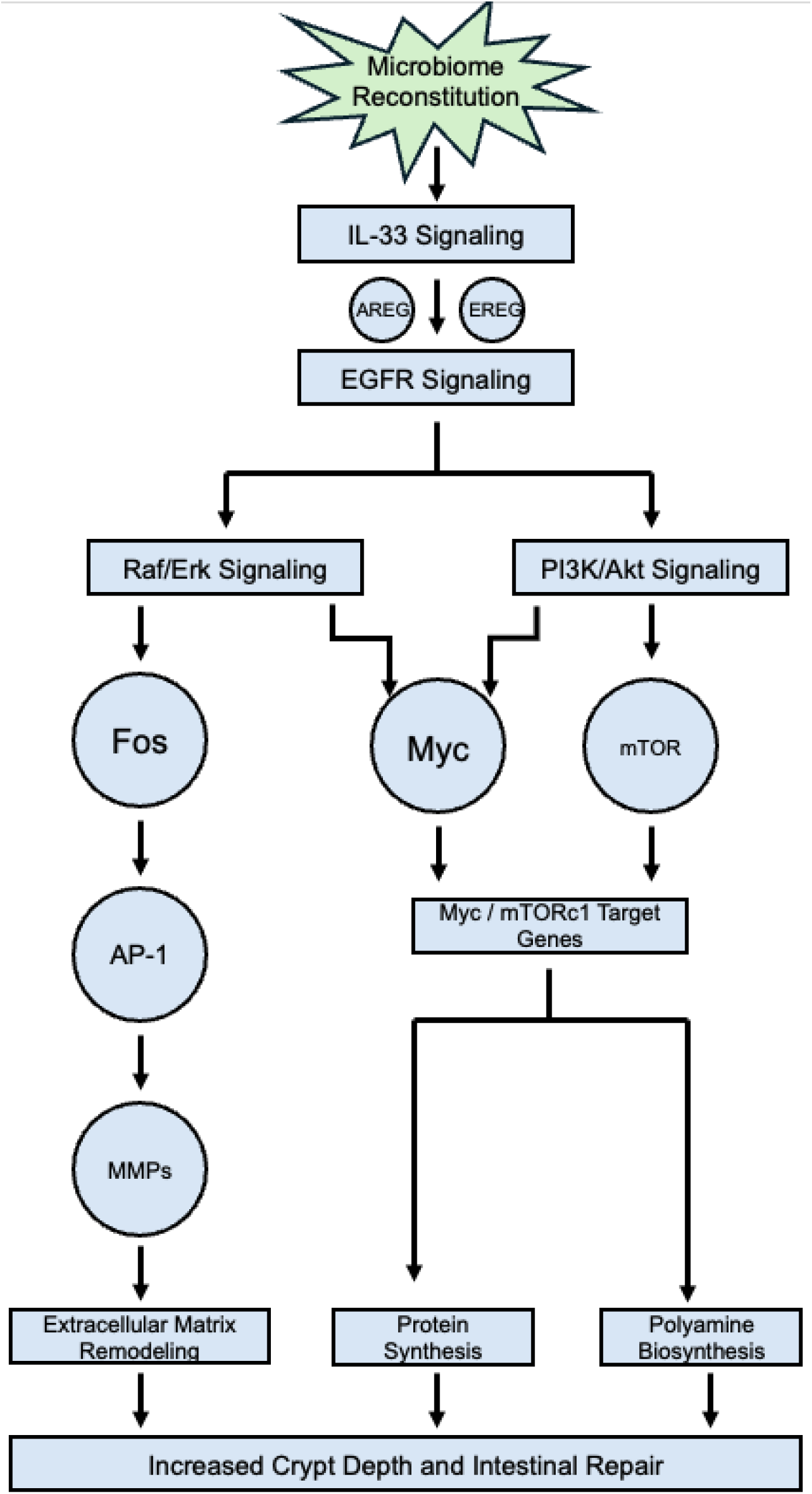
Visual summary of FMT-promoted changes in colonic gene expression. FMT is associated with significant increases in IL-33 signaling genes. IL-33 signaling promotes expression of EGFR ligands, including amphiregulin, which can stimulate Erk/MAPK and PI3K/Akt/mTOR signaling cascades. Erk and mTOR also stabilize Myc protein levels, promoting enhanced expression of Myc-activated genes. Myc and mTORc1 target genes promote ribosome biogenesis, protein synthesis and processing, and biosynthesis of polyamines, which promote proliferation of intestinal epithelial cells. Meanwhile, Erk signaling activates c-Fos, a component of the AP-1 transcription factor that increases matrix metalloprotease expression and promotes remodeling and repair of the extracellular matrix.

Further research is necessary to confirm the mechanistic links between these pathways and the changes in cell proliferation and crypt length observed in our histological data. However, these data suggest that FMT promotes regeneration of the intestine within a cohort of patients with recurrent CDI, and that this is associated with broad transcriptional changes in pathways such as Myc, mTOR, IL-33, and EGFR signaling.

## Data Availability

All data produced in the present study are available upon reasonable request to the authors

## Acknowledgements

We would like to thank the subjects in the FMT clinical trial for their cooperation and participation in this study. This work was supported by NIH grants R01 AI152477 and R01 AI124214 and the Henske Family. We are indebted to Drs. Jashim Uddin, Farha Naz, Mayuresh Abhyankar and Chelsea Marie for excellent discussion and review of this manuscript.

## Conflicts of Interest

WAP is a consultant for TechLab, Inc. that produces diagnostics for *C. difficile.* The other authors report no conflicts of interest.

## Notes

### Clinical Trial

NCT02797288

### Author Declarations

The Institutional Review Board of the University of Virginia gave ethical approval for this work.

